# The impact of reactive case detection on malaria transmission in Zanzibar in the presence of human mobility

**DOI:** 10.1101/2022.03.29.22273100

**Authors:** Aatreyee M. Das, Manuel W. Hetzel, Joshua O. Yukich, Logan Stuck, Bakar S. Fakih, Abdul-wahid H. Al-mafazy, Abdullah Ali, Nakul Chitnis

## Abstract

Malaria persists at low levels on Zanzibar despite the use of vector control and case management. We use a metapopulation model to investigate the role of human mobility in malaria persistence on Zanzibar, and the impact of reactive case detection. The model was parameterized using survey data on malaria prevalence, reactive case detection, and travel history. We find that in the absence of imported cases from mainland Tanzania, malaria would likely cease to persist on Zanzibar. We also investigate potential intervention scenarios that may lead to elimination, especially through changes to reactive case detection. While we find that some additional cases are removed by reactive case detection, a large proportion of cases are missed due to many infections having a low parasite density that go undetected by rapid diagnostic tests, a low rate of those infected with malaria seeking treatment, and a low rate of follow up at the household level of malaria cases detected at health facilities. While improvements in reactive case detection would lead to a reduction in malaria prevalence, none of the intervention scenarios tested here were sufficient to reach elimination. Imported cases need to be treated to have a substantial impact on prevalence.

## 1 Introduction

Despite a global reduction in malaria burden in 2000–2015, improvements in case incidence have stagnated in the United Republic of Tanzania at around 6 million cases per year since 2010 [1]. Zanzibar, a semi-autonomous region of Tanzania, has seen a substantial decline in malaria prevalence since 2000 due to the use of long-lasting insecticidal nets (LLINs), indoor residual spraying (IRS) and artemisinin-based combination therapies (ACTs) [2]. These strategies have aided in reducing malaria prevalence by 10- to 23-fold as measured by rapid diagnostic tests (RDTs) and microscopy, with prevalence estimated to be below 5% [2, 3] on both main islands of Zanzibar: Unguja and Pemba.

Additionally, the Zanzibar Malaria Elimination Programme (ZAMEP) has implemented a reactive case detection (RCD) programme from 2012 onwards [4]. RCD involves following up clinical malaria cases that present at a health facility and testing their household members for malaria using RDTs. This helps to treat both asymptomatic cases, and symptomatic cases that may not report to a health facility, with the aim to reduce onward transmission. RCD has been implemented with varying levels of success in countries and regions with low malaria prevalence such as China [5], Eswatini [6, 7], India [8], and Zambia [9]. ZAMEP was aiming to achieve follow up for 100% of confirmed cases by 2018 [10], but analyses of health facility data suggests that only 35.3% of diagnosed cases are followed up at the household level within 3 days [4].

Despite these substantial efforts, elimination has not been achieved in Zanzibar. The persistence of a low level of transmission despite high coverage of interventions has been attributed to geographic foci of transmission, a reservoir of sub-patent infections that are not detected and eliminated by routine surveillance-response activities, and repeated importation of infections [2]. The impact of these factors on disease transmission can be studied through mathematical modelling. Failing to account for these factors when modelling the disease can lead to overly optimistic estimates of the time or resources needed to eliminate malaria from a setting [2, 11].

Previous studies of RCD in Zambia and Namibia have suggested that it will only lead to malaria elimination in limited settings, particularly in areas that have reduced transmission recently [12, 13, 14, 15, 16]. The effectiveness of RCD can be improved by shifting to a reactive focal mass drug administration (rfMDA) programme, so that the probability of treating an infection is not dependent on the diagnostic test sensitivity [15, 16]. Diagnostic test sensitivity has been identified as a major impediment to RCD programmes in various settings, including Zanzibar [3], Zambia [9], and Eswatini [7], due to a high prevalence of very low density infections. Additionally, it has previously been suggested that RCD may not be useful in areas seeing large numbers of imported cases, as RCD relies on clusters of cases arising from local transmission [17]. On the other hand, as members of households often travel together, there is evidence that testing among the co-travellers of imported cases has a higher likelihood of yielding positive results [3]. Thus, it is unclear how useful RCD may be in the face of ongoing importation.

Previous studies of malaria importation have examined the impact of continuous importation of cases to Zanzibar from mainland Tanzania, where malaria prevalence is substantially higher [11, 18, 19, 20]. Parasite importation has also been shown to be an important factor for the persistence of malaria in settings outside of Tanzania [21, 22]. Churcher *et al* (2014) use branching process theory to calculate the reproduction number based on the proportion of detected cases that are classed as imported cases. If greater than 50% of detected cases are imported cases, the area is said to have a reproduction number below 1 and thus have halted endemic transmission [21], that is, indigenous incidence of malaria infection would not persist if all importation were halted [23]. Estimates of the proportion of clinical malaria patients in Zanzibar with a recent history of travel to mainland Tanzania have ranged from 9% to 49% [2, 3]. Whole genome sequencing of isolates from Zanzibar and mainland Tanzania has also highlighted the close relatedness of *Plasmodium falciparum* strains on Zanzibar and coastal Tanzania, suggesting some cases on Zanzibar have a recent history of importation [19].

A modelling analysis of malaria importation on Zanzibar has previously been conducted using mobile phone data to track human movement to and from mainland Tanzania [18, 24]. Using call data from the busiest period of travel to and from Zanzibar in 2008, Le Menach *et al* [18] estimated around 1.6 (falling within a range of 0–3.7) cases were imported per 1000 people per year to Zanzibar. The controlled reproductive number, *R*_*c*_, is the expected number of secondary human infections stemming from one untreated infection in an area with vector control measures in place. *R*_*c*_ was estimated to be within 0—0.56 in urban Unguja, 0.71—0.91 in rural Unguja, and 0.92—0.98 in rural Pemba, using an adapted Ross-Macdonald model. Another study looking at quantifying migration across country borders in East Africa using census data suggests that the majority of case importation in Tanzania from long-term migration is likely to occur near country borders, away from Dar es Salaam and Zanzibar [25]. To the best of our knowledge, no modelling study has yet been conducted on quantifying the impact of reactive case detection on malaria transmission in Zanzibar, particularly in the presence of ongoing human movement and case importation rates that change in line with changes in prevalence in other areas.

In this paper, we use a compartmental metapopulation model to examine the impact of RCD and rfMDA, combined with ongoing short-term human movement, on the persistence of malaria in Zanzibar and the potential impact of treating imported cases. Using malaria prevalence estimates for the islands of Pemba, Unguja and mainland Tanzania, along with data on the RCD programme, we consider the potential effects of improving or reducing the RCD programme currently in place, including changes in follow up, improvements in the number of cases reporting to health facilities, additional testing of neighbours of index cases, and shifting to an rfMDA intervention. We also consider possible synergies to be gained by combining rfMDA with treating neighbours as well as index households. Finally, combining the malaria prevalence estimates with travel history data, we estimate the likely impact of treating a proportion of imported infections on malaria prevalence on Zanzibar.

## 2 Methods

This analysis uses two main models: a compartmental susceptible-infected-susceptible (SIS) population model that was adapted to describe transmission dynamics in the presence of short-term human movement, and a stochastic implementation of this model including an ongoing RCD programme in Zanzibar. The first model is used to understand the role played by human movement in the persistence of malaria on the islands, and the second model is used to understand the impact of interventions strategies such as RCD and the treatment of imported cases in reducing the endemic equilibrium on the islands. Both models consisted of three patches: Pemba, Unguja, and mainland Tanzania.

### 2.1 Study setting

The Zanzibar archipelago lies to the east of the mainland of the United Republic of Tanzania. According to the 2012 census, the two main islands, Unguja and Pemba, had populations of 896,721 and 406,848, respectively. The islands are connected to mainland Tanzania via ferries and two airports (Fig. 1). In addition to this, there is regular small boat traffic between mainland Tanzania and Zanzibar, often by traditional dhows.

**Figure 1:**
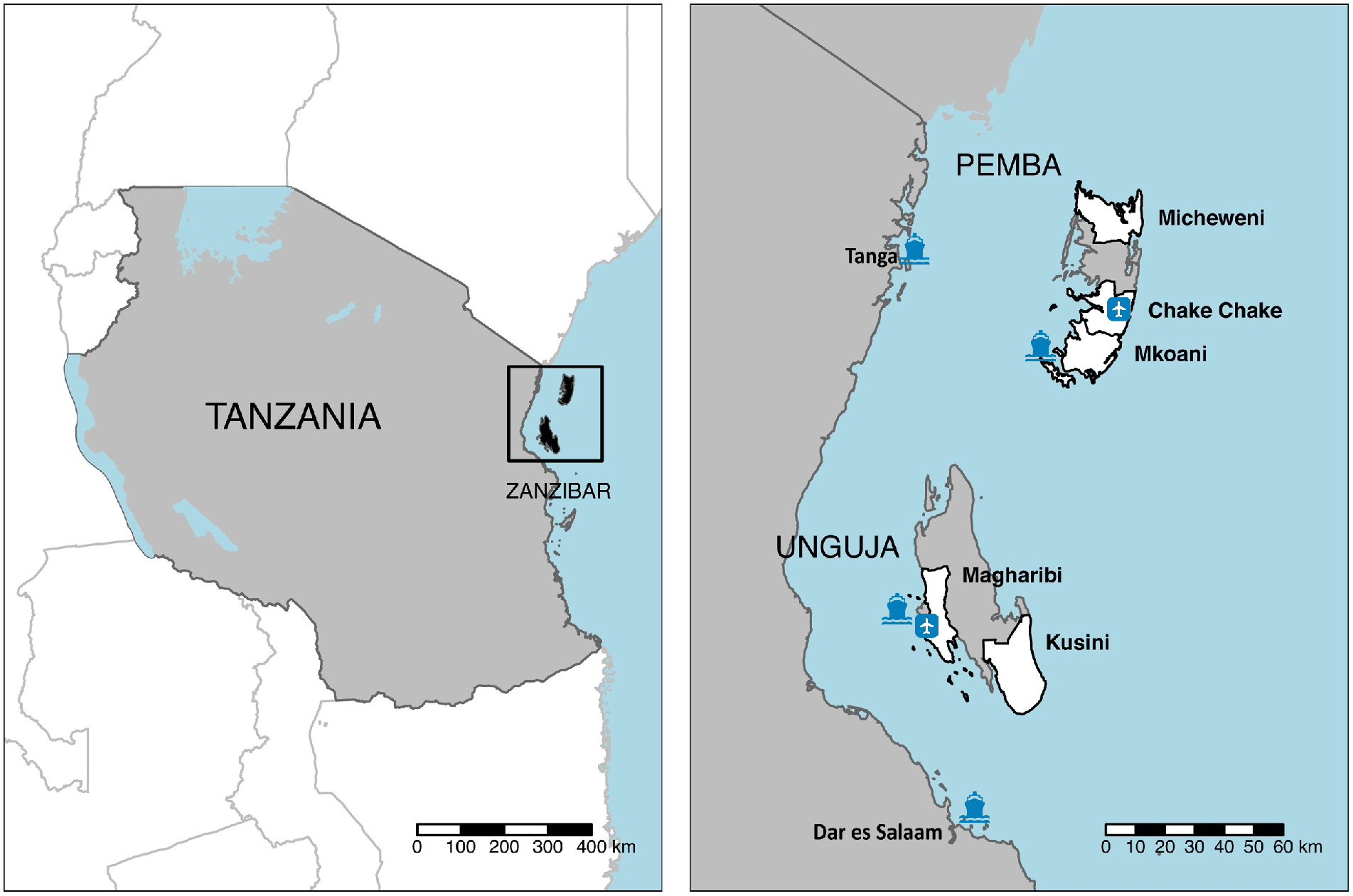
Map of Zanzibar, with the RADZEC study districts in white. Airports and ferry terminals are highlighted. Figure adapted from [3].

ZAMEP runs an RCD programme to effectively target test-and-treat efforts towards foci of infection. When patients on either island are diagnosed with malaria at a health facility, they should ideally be followed up within 3 days at their household by a district malaria surveillance officer (DMSO), and all household members should be tested with an RDT for malaria. Those who return a positive test result are treated with artesunate-amodiaquine and a single dose of primaquine. The Reactive Case Detection in Zanzibar: System Effectiveness and Cost (RADZEC) study included an examination of the operational coverage of the RCD programme [4]. Across the 150 public health facilities and 51 private health facilities, a mean of 32 and 12 malaria cases arrived at a health facility per district per month in Unguja and Pemba, respectively, corresponding to 6.4 cases per day in the whole of Unguja, and 1.6 in Pemba. Of those diagnosed at a health facility, 35.3% were followed up at the household by a district malaria surveillance officer within 3 days, 47.9% within 6 days, 59.9% within 15 days, and 62.0% within 21 days. The mean household size for index households was found to be 7.0 people per household on Pemba and 6.2 people per household on Unguja, including index cases [3].

This data, along with rolling cross-sectional survey data from the RADZEC study, were used to parameterise the RCD parameters in the model.

### 2.2 RADZEC cross-sectional survey data

The rolling cross-sectional survey component of the RADZEC study was conducted between May 2017 and October 2018. It involved following DMSOs on visits to the households of patients diagnosed with malaria at a health facility (from now on referred to as the index case). A cross-sectional survey was conducted at these households, which included a questionnaire, RDT tests, and collecting blood samples for quantitative polymerase chain reaction (qPCR) tests. The survey included three types of households: index case households, neighbouring households, and a transect of households stemming from the index household. Neighbouring households consisted of the four households nearest to the index case household, and transect households consisted of five households along a 200m transect starting from the index household. The full survey details are described elsewhere [3].

The survey collected data on a range of factors including demographics, a recent history of illness, and detailed travel history from the last 60 days. The median trip length was found to be six nights.

Within the survey population, 12,487 residents were tested with RDTs for malaria and 6,281 with qPCR tests. The sensitivity of RDTs to detect qPCR-detectable infections was found to be 34% [3].

The malaria prevalence on each island was estimated by first taking the number of PCR-positive test results outside of the index household above a cut-off of 0.13 parasites/*μ*l, below which the chance of false positive results increases. The number of people with PCR-positive results in neighbouring and transect households was divided by the total number of people tested in neighbouring and transect households on each island to give the estimated prevalence on each island. Members of the index household were not included as this would have led to an artificial inflation of the malaria prevalence as index households contained a known malaria case (the index case) and had a higher likelihood of containing additional cases [3]. As this data was collected around the households of index cases, there was a possibility that the prevalence in this sample was still higher than in a random sample. At the same time, as this method directly excludes index cases and index households, where malaria prevalence is typically higher, there was a chance that the prevalence found in neighbouring and transect households would be an underestimate. In order to compare to a random sample, the qPCR prevalence in neighbouring and transect households in Micheweni district (north Pemba) in the RADZEC dataset was compared to the mean prevalence found by qPCR in a randomly sampled cross-sectional survey conducted in Micheweni in 2015 [2]. The prevalence in neighbouring and transect households in the RADZEC study was 1.8% (95% CI: 0.9-2.7), while the prevalence in the cross-sectional survey conducted in a random sample of households was 1.7% (95% CI: 1.1–2.4). This suggests that the positivity rate in neighbouring and transect households is a good approximation of the population prevalence.

The mean number of neighbours tested per index case was 20.4 in Pemba and 18.2 in Unguja. The ratio of cases amongst index household members compared to neighbouring and transect households was 3.2 in Pemba and 10.0 in Unguja. The ratio of cases in neighbouring households compared to neighbouring and transect households was 0.8 in Pemba and 1.3 in Unguja.

Travel data suggested that travellers spend similar numbers of nights in multiple parts of mainland Tanzania, so the malaria prevalence for 2-10 year old children across all of Tanzania, as estimated by the Malaria Atlas Project, was taken as the baseline for mainland Tanzania [26]. This is likely an overestimate of the population prevalence, as the prevalence in 2-10 year old children is typically higher than in the general population [27].

Time spent away from home, captured in the travel matrix *θ*_*ij*_ (see Table 1), was calculated by noting which proportion of nights in the last 60 nights were spent away from home amongst survey respondents from each patch and where they were spent, where *i* and *j* represent Pemba, Unguja and mainland Tanzania,

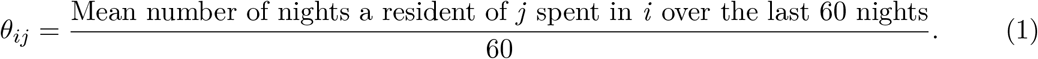

where 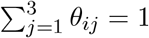.

**Table 1:** Descriptions of state variables and parameters for the SIS model with human movement.

| Parameter or state variable | Description and units |
| --- | --- |
| $I_k$ | Proportion of people who are infectious in patch $k$. Dimensionless. |
| $I_k^*$ | Proportion of people who are infectious in patch $k$ at equilibrium, before changes to ongoing interventions are applied. Dimensionless. |
| $N_k$ | Total number of people in patch $k$. Humans. |
| $\beta_k$ | The effective malaria transmission rate from humans to other humans in patch $k$. Day<sup>-1</sup>. |
| $\theta_{ij}$ | The proportion of time the average resident of patch $j$ spends in patch $i$. Dimensionless. |
| $\mu$ | Natural infection clearance rate. Day<sup>-1</sup>. |

As we did not have data on travel to Zanzibar by residents of mainland Tanzania, we have assumed that the same number of person-nights are spent in total by residents of mainland Tanzania on Zanzibar as the other way around. Thus,

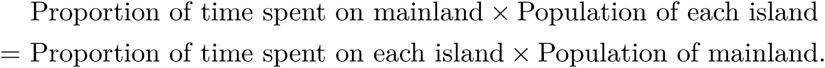

Treatment was not included in the models outside of treatment due to RCD (which also includes treatment of the index case). Instead, the daily natural clearance rate was taken to be (1*/*200)day^−1^ [28, 29].

### 2.3 Model description

Human movement was modelled using a deterministic SIS metapopulation model including three patches for Pemba, Unguja and mainland Tanzania. This was then extended to include stochasticity and the effects of RCD on Pemba and Unguja.

A model schematic can be found in Fig. 2.

**Figure 2:**
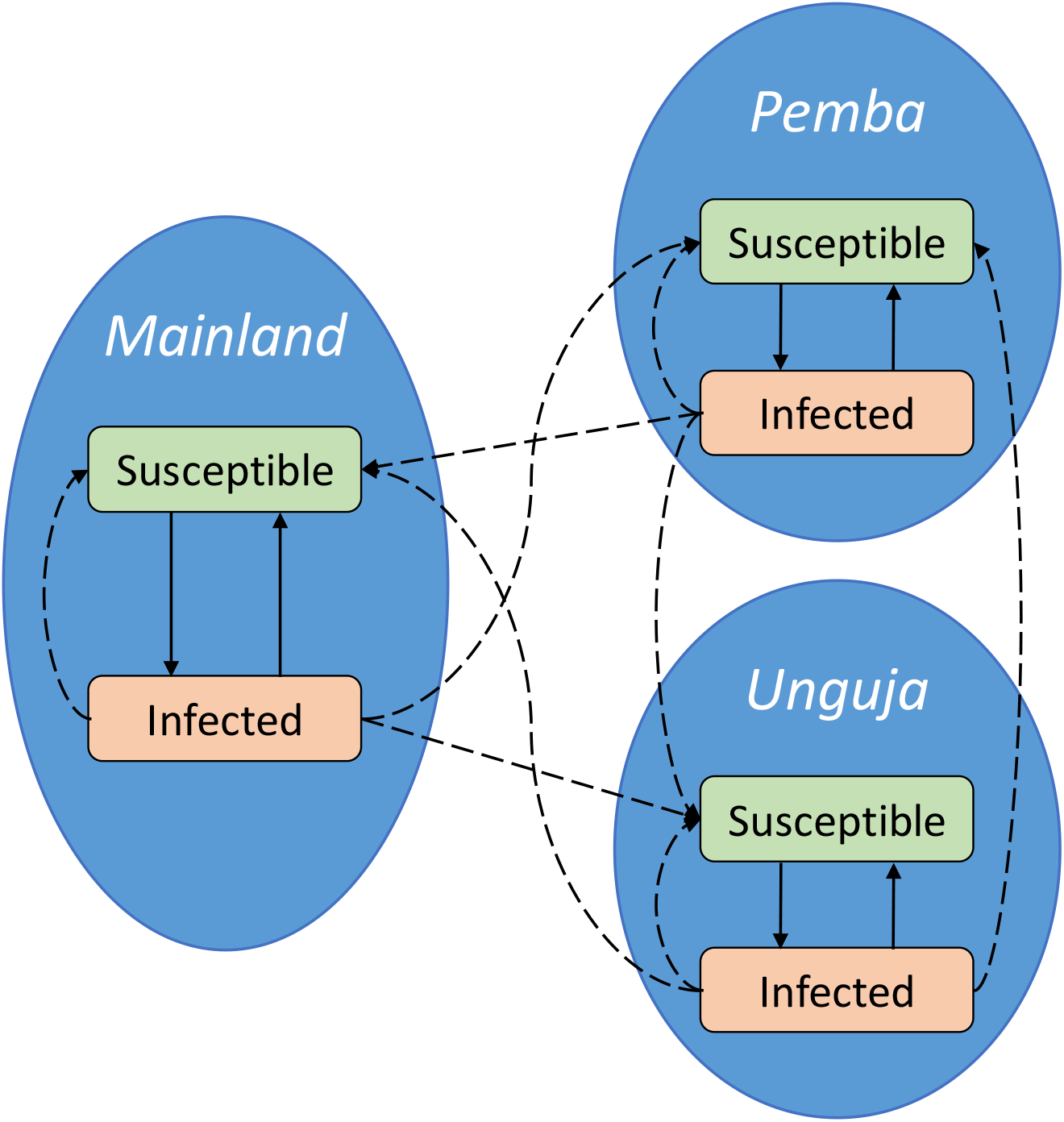
A schematic diagram of the model with two disease states in each patch. Solid arrows represent transitions between disease states, and dashed arrows represent transmission.

#### 2.3.1 SIS model including human movement

The total number of people from patch *j* spending time in patch *i*, weighted by the amount of time they spend there, is given by *N*_*j*_*θ*_*ij*_. Similarly, *N*_*j*_*θ*_*ij*_*I*_*j*_ gives the number of infected people from patch *j* spending time in patch *i*, weighted by the amount of time they spend there. When combined, the effective proportion of the population that is infectious in patch *i* is given by

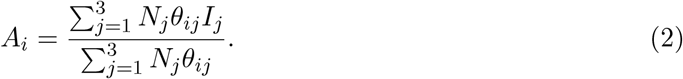

A description of the parameters and state variables can be found in Table 1.

*β*_*i*_*A*_*i*_*θ*_*ik*_ is the contact rate between a susceptible individual from patch *k* and an infected individual in patch *i*. Summing over *i* gives the total rate at which a susceptible individual in patch *k* comes into contact with an infected person either in their own patch or another patch, and becomes infected,

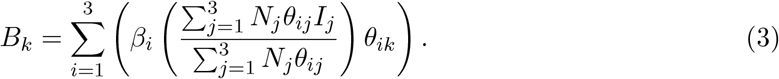

Eq. (3) is adapted from previous work by Ruktanonchai *et al* [30], accounting for both the infectious people moving in and out of patch *k*, as well as the time spent by residents of *k* in other patches.

Combining this with the proportion of susceptible individuals in patch *k*, which we represent as *S*_*k*_ = 1 − *I*_*k*_, and allowing infected individuals to recover at the natural clearance rate of the disease gives the overall equation for the rate of change of *I*_*k*_:

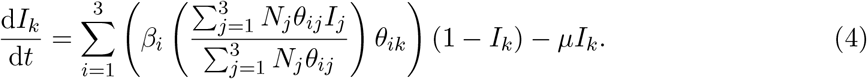

We assume that the majority of trips are short-term trips and people retain the properties of their residential patch in terms of recovery rate and, in section 2.3.2, the RCD programme. Survey responses about travels in the last 60 days support the assumption of short trips.

We calibrated the model by assuming that malaria prevalence is at equilibrium. Under this assumption, we can calculate the transmission rate that would lead to the observed prevalence. Thus, setting the right hand side of Eq. (4) to 0,

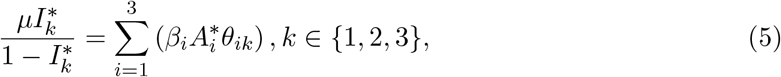

where 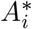 is *A*_*i*_ at the equilibrium prevalence.

The transmission parameter, *β*, encompasses malaria transmission from humans to mosquitoes and back again, along with any malaria control strategies already in place. It can be derived as the solution to a set of simultaneous equations:

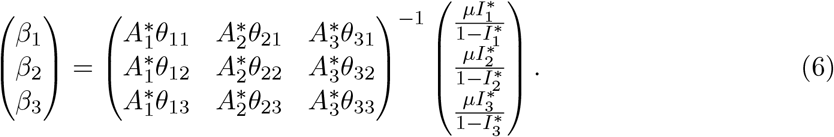

We then used the deterministic model to estimate the impact of human movement on malaria persistence on the islands of Pemba and Unguja. The impact of no human movement was modelled by keeping the calibrated transmission and recovery rates constant, but changing the time spent away from the home patch to 0 in all cases (i.e. *θ*_*ij*_ = 0 for all *i* ≠ *j* and *θ*_*ij*_ = 1 for all *i* = *j* for *i, j* ∈ 1, 2, 3). This scenario acts as a counterfactual for deducing how human movement contributes to the persistence of malaria despite the current use of interventions.

#### 2.3.2 SIS model including human movement and an RCD programme

Eq. (4) is modified in line with previous work by Chitnis *et al* [12] to include RCD. RCD is modelled by removing a number of infected individuals proportional to the number of infected people in that patch. The rate of change in *I*_*k*_ is now given by

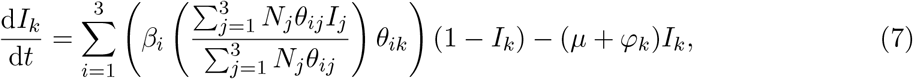

where *ϕ*_*k*_ is the rate of removing people from the infected class due to the RCD programme. This is the product of the number of cases followed up by the RCD programme per day, the mean number of household members in each index house, the ratio of positive tests in an index house versus the general population, and the test positivity rate, divided by the total population in that patch,

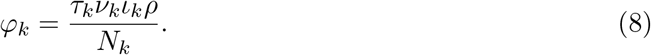

Parameter descriptions can be found in Table 2. The number of cases followed up by the RCD programme per day depends on the number of infected people at any given time, the rate of seeking treatment, and the proportion of cases followed up by DMSOs,

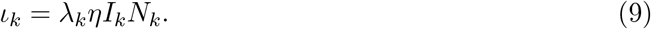

**Table 2:** Descriptions of RCD programme parameters.

| Parameter | Description and units |
| --- | --- |
| $\varphi_k$ | Treatment rate due to RCD programme in patch $k$. $\text{Day}^{-1}$. |
| $\varphi_k^*$ | Treatment rate due to RCD programme at equilibrium in patch $k$. $\text{Day}^{-1}$. |
| $\tau_k$ | Ratio of malaria prevalence in individuals tested within the RCD programme as compared to the general population in patch $k$. Dimensionless. |
| $\iota_k$ | Number of cases followed up by a District Malaria Surveillance Officer per day in patch $k$. Humans per day. |
| $\iota_k^*$ | Number of cases followed up by a District Malaria Surveillance Officer per day at equilibrium in patch $k$. Humans per day. |
| $\nu_k$ | Number of people tested during follow up per index case in patch $k$. Dimensionless. |
| $\rho$ | Rapid diagnostic test sensitivity. Dimensionless. |
| $\lambda_k$ | The daily rate at which an infected individual seeks treatment in patch $k$. $\text{Day}^{-1}$. |
| $\eta$ | The proportion of cases arriving at the health facility that are followed up by the DMSO. $\text{Day}^{-1}$. |
| $\eta^*$ | The proportion of cases arriving at the health facility that are followed up by the DMSO within 3 days. Dimensionless. |

The rate of seeking treatment is assumed to be constant and is calculated from the observed number of cases arriving at the health facility at equilibrium,

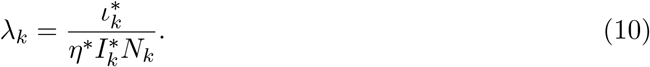

The baseline value for the proportion of cases followed up by a DMSO at the index case household level, *η*^*\**^, was taken to be the 3 day follow up rate: 35.3%.

Descriptions of RCD parameters can be found in Table 2.

We compared testing only index household members in the RCD programme and testing both the index household and neighbours. When considering just index households, the targeting ratio was calculated by taking the ratio of the positivity rate, as measured by PCR, in index households compared to neighbouring and transect households. This was then adjusted in the model to ensure that a positive case was included for the index case, as often the index case had been treated by the time the DMSO followed up the case at the index household. The targeting ratio, *τ* ^(*h*)^, given in Table 3 considers only the prevalence in index household members outside of the index case. When considering neighbouring households as well, the targeting ratio in neighbouring households was calculated by taking the ratio of PCR-positive cases in neighbouring households as compared to both neighbouring and transect households. We find that the likelihood of finding a case is 10 times higher in the index household than in a neighbouring household in Unguja, and 5 times higher in Pemba. The equation for *ϕ*_*k*_ was adapted to

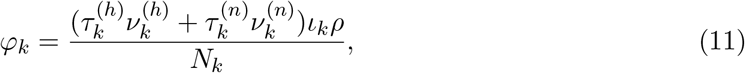

where the superscripts *h* and *n* refer to the index household and neighbouring households, respectively.

**Table 3:** Variable and parameter values and sources. Where a range of parameter values were tested in the sensitivity analysis, the 95% confidence interval for the range of values tested is given. For *θ*_*ij*_, the order of the rows and columns of the matrix correspond to Pemba, Unguja and mainland Tanzania.

| Variable or parameter | Mean values [95% CI] |  |  | Source |
| --- | --- | --- | --- | --- |
|  | Pemba | Unguja | Mainland |  |
| $I_k^*$ | 1.36%<br>[0.96-1.93] | 1.18%<br>[0.86-1.61] | 7.79% | [3, 26] |
| $N_k$ | 406,848 | 896,721 | 43,625,354 | [31] |
| $\theta_{ij}$ | $\begin{pmatrix} 0.991 & 0.004 & 5.7 \times 10^{-5} \\ 0.003 & 0.970 & 5.3 \times 10^{-4} \\ 0.006 & 0.026 & 0.999 \end{pmatrix}$ | | | [3] |
| $\mu$ | 0.005 day<sup>-1</sup> | 0.005 day<sup>-1</sup> | 0.005 day<sup>-1</sup> | [28, 29] |
| $\tau^{(h)}$ | 3.2 [2.0-4.8] | 10.0 [8.0-12.6] | N/A | [3] |
| $\tau^{(n)}$ | 0.7 [0.4-1.3] | 1.3 [0.9-1.9] | N/A | [3] |
| $\nu^{(h)}$ | 7.0 [6.5-7.5] | 6.3 [5.9-6.9] | N/A | [3] |
| $\nu^{(n)}$ | 20.4 [19.4-21.4] | 18.8 [17.6-19.9] | N/A | [3] |
| $\rho$ | 34% | 34% | N/A | [3] |
| $\eta^*$ | 35.3% | 35.3% | N/A | [4] |
| $\eta$ - range of values tested | 0%, 35%, 48%, 60%, 62%, 100% | 0%, 35%, 48%, 60%, 62%, 100% | N/A | Values based on DMSO follow up at the index case household level observed in [4] |

The RCD programme has been running on Zanzibar since 2012. We assume that the malaria prevalence has reached a steady state since the introduction of RCD. Case incidence data from 2012 to 2015 shows seasonal trends but relatively stable incidence over this time period [32]. Setting the right hand side of Eq. (7) to 0 and solving for *β* gives the transmission rates in the presence of an ongoing RCD programme,

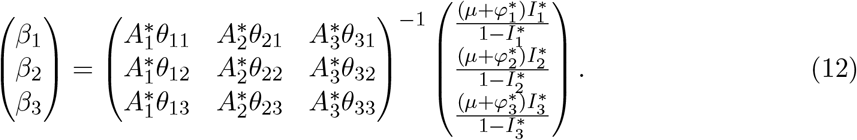

New interventions or potential changes to interventions are only simulated post-calibration. The transmission parameter on the three islands is unaffected by the new intervention, since all interventions considered here only target the infectious reservoir in humans and not the vectorial capacity.

#### 2.3.3 Treatment of imported cases

Currently prophylaxis is not given to travellers when travelling to mainland Tanzania or vice versa. Similarly, there is no screen-and-treat programme for entrants to Zanzibar. We expanded our model to include treatment of imported cases as a potential intervention, in order to evaluate what proportion of cases must be treated to achieve different reductions in prevalence on Pemba and Unguja. Eq. (7) was modified to have a *θ*^outbound^, which included treatment for mainland Tanzanians on their outbound journey to Zanzibar, and *θ*^return^ for Zanzibari residents that receive treatment on their return journey to Zanzibar. Thus Eq. (7) was modified to

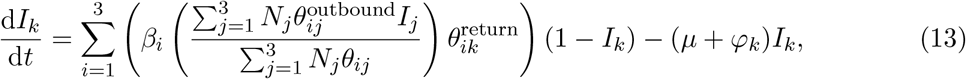

where

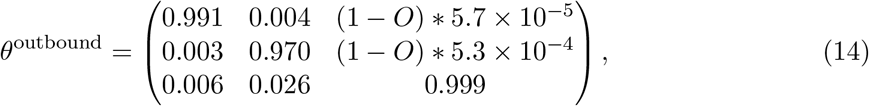

and

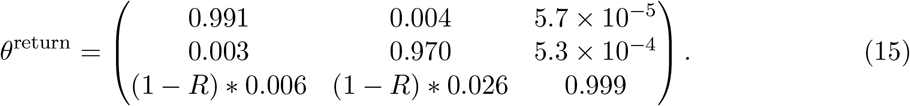

*O* represents the proportion of travellers from mainland Tanzania receiving treatment such that they are no longer infected upon entering Zanzibar, and *R* represents the proportion of Zanzibari residents receiving treatment such that they are no longer infected upon returning to Zanzibar.

### 2.4 Simulations

Stochastic simulations were only run with the model with RCD. In order to allow for small but finite populations of infectious individuals, a binomial tau-leap adaptation of the Gillespie algorithm was used to model Eq. (7) [33]. Following calibration, the current RCD programme (baseline of 35.3% follow up of index cases at index households only) was compared to a range of alternatives:

1. RCD at a range of levels of follow up (see Table 3 for values);
2. Expanding the RCD system to include follow up at four neighbouring households as well;
3. Doubling the daily treatment seeking rate;
4. rfMDA in the index household rather than test-and-treat;
5. Treating 50% of imported cases.

The effects of varying the proportion of cases followed up at the household level is tested by varying the follow up proportion between those seen in 3, 6, 15 and 21 days, as well as stopping the RCD programme altogether (no follow up) and perfectly following up every case. The potential benefits of testing and treating all neighbours in approximately four nearby households as well was considered. As the rate of seeking treatment amongst those infected is low, we tested doubling the daily treatment seeking rate (e.g. by promoting early treatment seeking or broader testing of patients at formal health facilities, or due to more individuals being symptomatic due to waning immunity). Additionally, rfMDA was modelled with the same parameters as for RCD, except the value of the test sensitivity was changed to 100%, as all index household members, infected or susceptible, would automatically receive treatment. Finally, treating 50% of cases imported onto the islands by either Zanzibari residents travelling to mainland Tanzania, or visitors from mainland Tanzania were also modelled (*O* = *R* = 0.5). 500 simulations were run for each combination of intervention parameters.

### 2.5 Impact of parameter uncertainty

The impact of parameter uncertainty was investigated by testing a range of parameter values in a sensitivity analysis. The values were based on the uncertainty in the sample data. The parameters varied and the distributions from which they were sampled were as follows:

- The equilibrium malaria prevalence on Pemba, 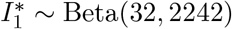
- The equilibrium malaria prevalence on Unguja, 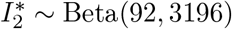
- The targeting ratio in index households in Pemba, 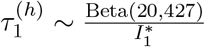
- The targeting ratio in index households in Unguja, 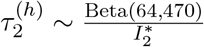
- The targeting ratio in neighbouring households in Pemba, 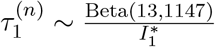
- The targeting ratio in neighbouring households in Unguja, 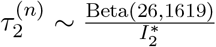
- The number of people tested by the RCD programme in the index household in Pemba, 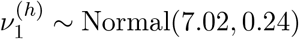;
- The absolute number of people tested by the RCD programme in the index household in Unguja, 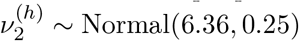;
- The absolute number of people tested by the RCD programme in neighbouring households in Pemba, 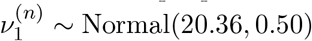;
- The absolute number of people tested by the RCD programme in neighbouring households in Unguja, 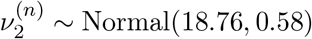.

Subscripts of *1* and *2* indicate Pemba and Unguja, respectively. 100 random values were selected from these parameter distributions, and each set of values was simulated with five different seeds, forming a total of 500 simulations for each intervention. The 95% confidence intervals of the distributions used for these parameters can be found in Table 3.

## 3 Results

### 3.1 SIS model including human movement

The SIS transmission model described by Eq. (4) showed standard dynamics of reaching the equilibrium prevalence seen in the RADZEC study. When human movement was removed by changing the movement matrix, *θ*, to an identity matrix, the equilibrium prevalence dropped to zero on both Pemba and Unguja. This result is to be expected, as the calibrated transmission parameter is lower than the natural parasite clearance rate in both Pemba and Unguja. The calibrated values for *β* were 0.0048 (95% CI: 0.0044-0.0050), 0.0037 (95% CI: 0.0025-0.0.047) for Pemba and Unguja, respectively. *R*_*c*_, given by the transmission rate divided by the recovery rate, was found to be 0.95 (95% CI: 0.88-1.00) on Pemba and 0.74 (95% CI: 0.50-0.94) on Unguja.

An analysis of the reproductive number of the whole system showed that the overall reproductive number is highly dependent on the transmission rate on mainland Tanzania. Details of this can be found in the Supplementary Information.

### 3.2 SIS model including human movement and an RCD programme

All simulations were initially calibrated to the baseline scenario of 35.3% follow up of index cases at the household level only. Year 0 is when the intervention is introduced.

Fig. 3 shows the timeseries expected from removing RCD that is currently in place. The proportion of index cases followed up by a DMSO was set to 0 from year 0. We observe a rise in the malaria prevalence until a new equilibrium is reached. The 50% and 95% confidence intervals of the 500 simulations at each time point are also included, alongside the median number of infected individuals. For illustration purposes, three individual stochastic simulations are also included to show how the malaria prevalence may vary within a single simulation. While individual simulations can fluctuate quite a lot, the median settles to a pseudo-equilibrium. We estimate that removing RCD would lead to a 10% increase in malaria prevalence on Pemba, and an 8% increase in prevalence on Unguja.

**Figure 3:**
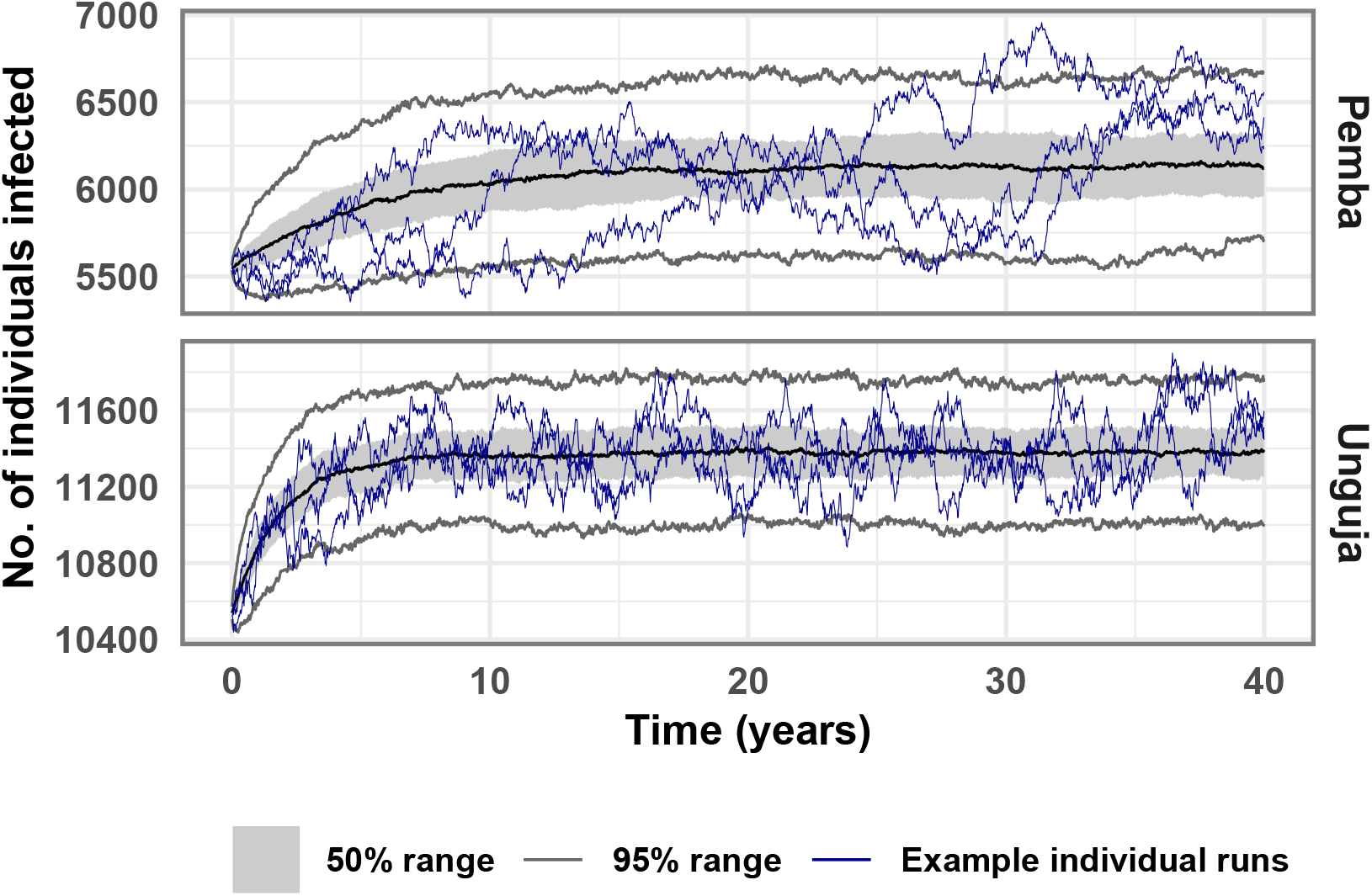
Timeseries of 500 stochastic simulations showing the median, 50% confidence interval, 95% confidence interval, and three individual simulation runs, for the scenario where all RCD is stopped at year 0.

Fig. 4 shows the impact of increasing the proportion of cases followed up by a DMSO in a timely manner from the 3-day follow up proportion of 35%, to the 21-day follow up proportion of 62%, and then to 100%. The final malaria prevalence reached under these intervention scenarios are compared to the baseline malaria prevalence (RCD with 35% of index cases followed up) and a counterfactual which indicates the pseudo-equilibrium reached when RCD is stopped (Fig. 3). Increasing follow up with no other changes to RCD has a very small effect on the final malaria prevalence reached after 40 years. Including 4 neighbouring households in RCD makes a negligible difference to the malaria prevalence. Shifting to rfMDA leads to some additional cases being treated due to the removal of testing. This decrease in prevalence is further compounded when combined with following up all index cases at the index household. Once again, including neighbouring households in rfMDA does not make a substantial difference. Doubling the rate at which infected people seek treatment and are identified as index cases leads to RCD finding and treating roughly twice as many cases. Finally, treating 50% of imported cases such that they cannot lead to further cases on Zanzibar led to large reductions in prevalence, with a 43% reduction in prevalence on Pemba and a 47% reduction in prevalence on Unguja.

**Figure 4:**
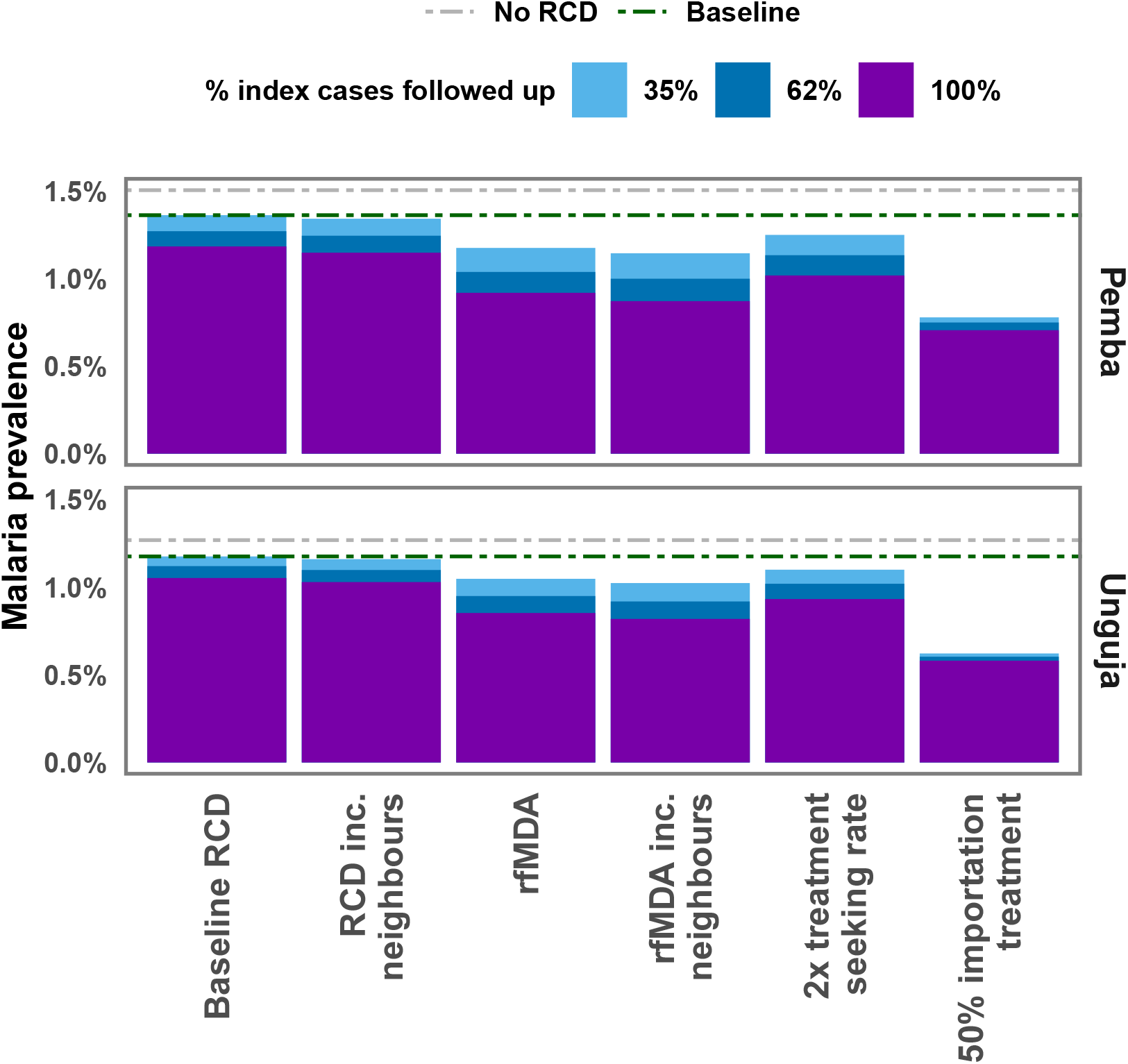
Median malaria prevalence reached after 40 years of simulations under different intervention scenarios and with different levels of follow up of index cases arriving at a health facility. Dashed lines indicate the baseline prevalence and the prevalence expected with no RCD. ‘Baseline’ refers to the malaria prevalence in the presence of RCD with 35% follow up of index cases, as observed in the RADZEC study.

Treating people who travel would need to achieve high coverage for both travellers to and from mainland Tanzania to achieve a substantial reduction in prevalence, as illustrated in Fig. 5. Time-series plots for a range of treatment proportions can be found in the Supplementary Information. Due to the transmission rate being substantially higher on Pemba than Unguja, even treating all malaria importations from mainland Tanzania would likely be insufficient to lead to elimination within 40 years on either Pemba or Unguja, as infections would be imported from Pemba to Unguja, sustaining transmission.

**Figure 5:**
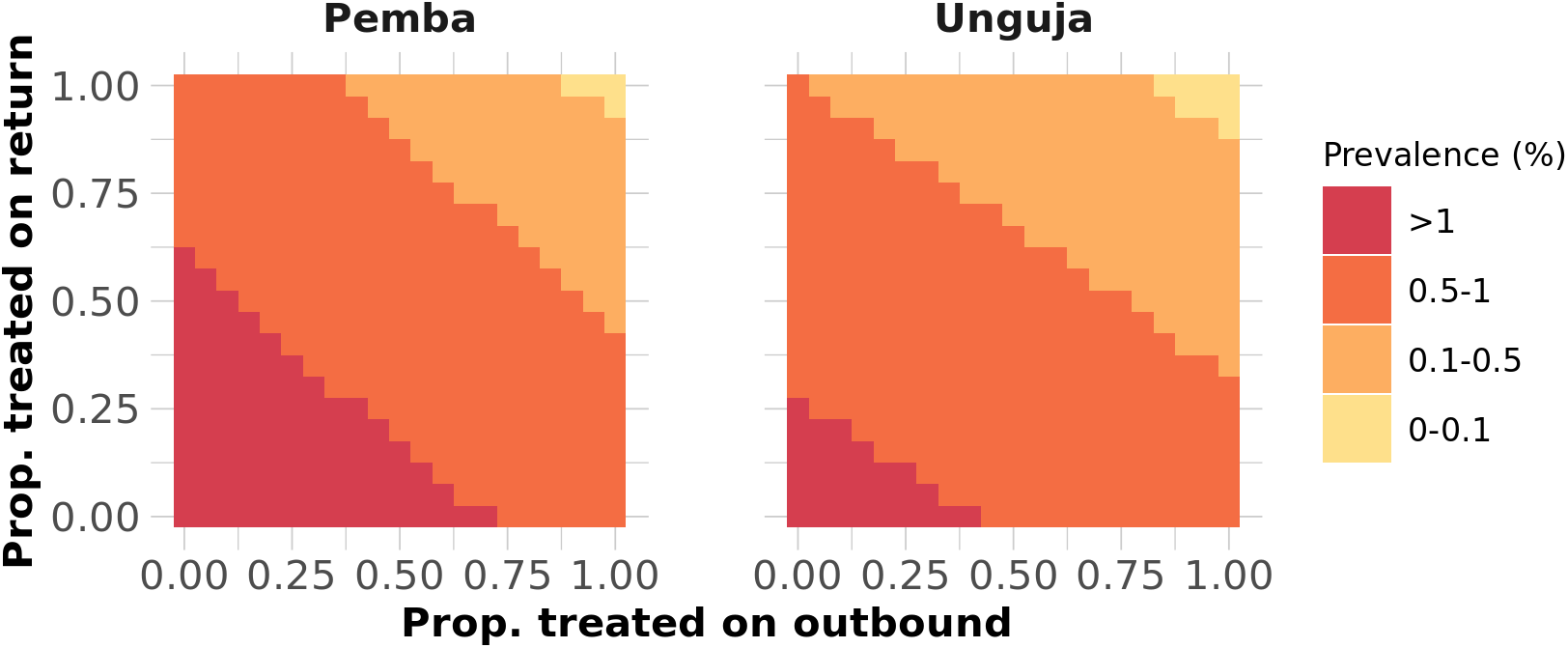
Heatmap showing the median final prevalence reached after 40 years out of 500 stochastic runs when treatment of travellers is included.

Fig. 6 shows the amount of resources needed for RCD and rfMDA when including or not including neighbours, or when following up 100% of cases at the index household level. This figure does not consider the RDTs or ACTs needed outside of RCD (e.g. RDTs used to detect index cases in the health facility or ACTs distributed through pharmacies for malaria treatment outside of RCD). It also does not consider the additional personnel and time needed to expand RCD to include neighbours. In general, including neighbours leads to a much larger use of resources but with little gains in malaria prevalence reduction.

**Figure 6:**
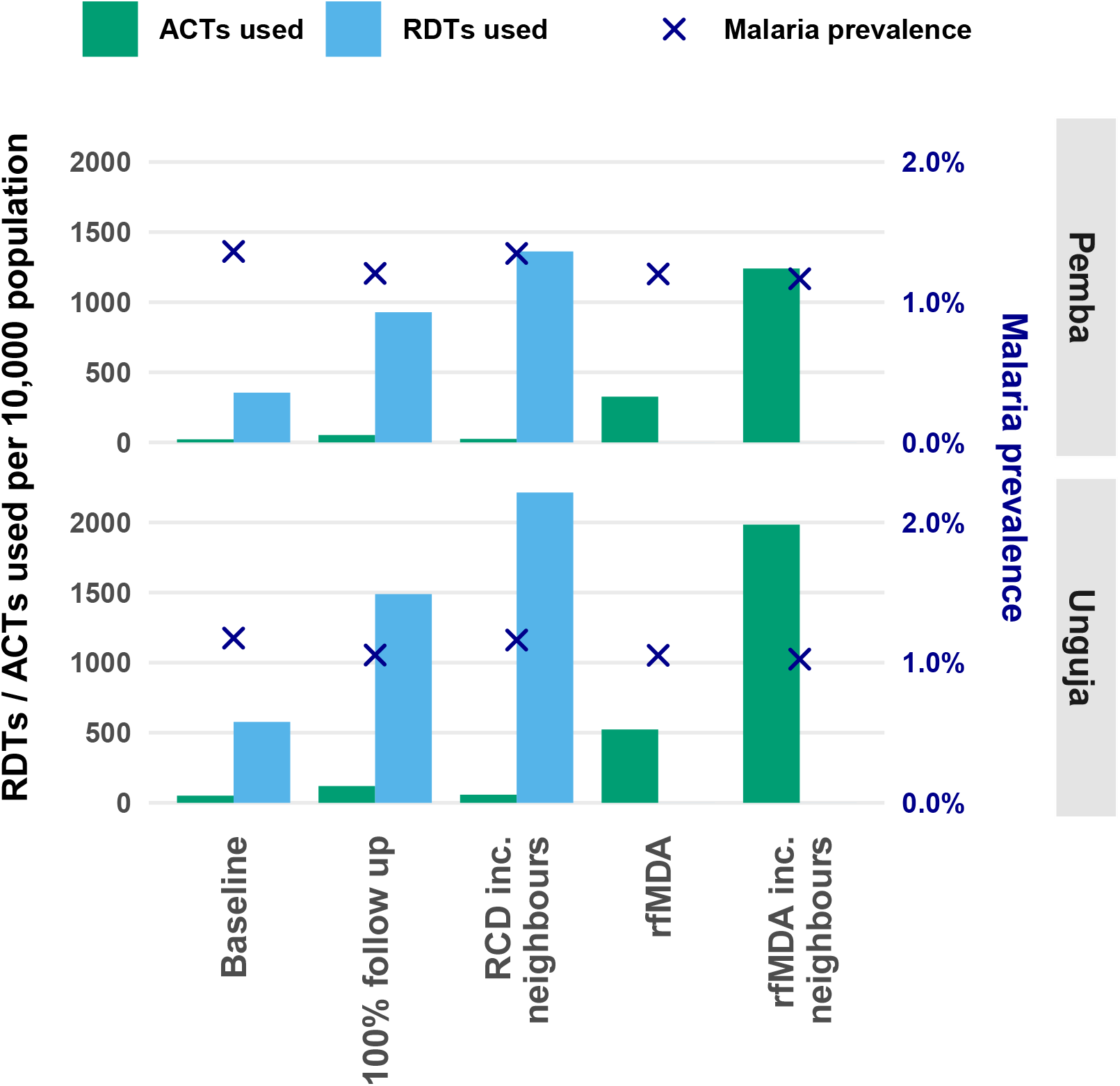
The cumulative number of RDTs and ACTs used per 10,000 population over 10 years since the start of interventions. Additionally, the crosses represent the malaria prevalence reached with that intervention. ‘Baseline’ refers to RCD with 35% follow up of index cases. ‘100% follow up’ refers to RCD with 100% follow up of index cases. ‘RCD inc. neighbours’ refers to RCD with 35% follow up of index cases and testing and treatment at the index household and four neighbouring households. ‘rfMDA’ refers to 35% follow up and presumptive treatment of index household members. ‘rfMDA inc. neighbours’ refers to following up 35% of index cases and presumptive treatment of the index household and members of four neighbouring households. Note, this does not include RDTs used for diagnosing index cases, or ACTs used in malaria treatment outside of RCD.

### 3.3 Impact of parameter uncertainty

Our analysis suggests that switching from RCD to rfMDA (at the same proportion of index cases followed up at the household level: 35.3%) has a similar impact as increasing the follow up proportion in the RCD programme to 100%, but neither increase the recovery rate sufficiently to lead to elimination. A larger decrease in prevalence is seen if rfMDA is implemented with 100% of follow up at the index case household level, or if 50% of imported cases are treated. When parameter uncertainty is included in the simulations, we find that although the final prevalence reached in 40 years is sensitive to the varied parameters (see Fig. 7), the overall trends remained the same. It is worth noting that the median prevalence reached after 40 years across 100 parameter sets differs from the prevalence reached in previous figures due to the difference in the median and modal values of the parameter distributions (see Supplementary Information for details).

**Figure 7:**
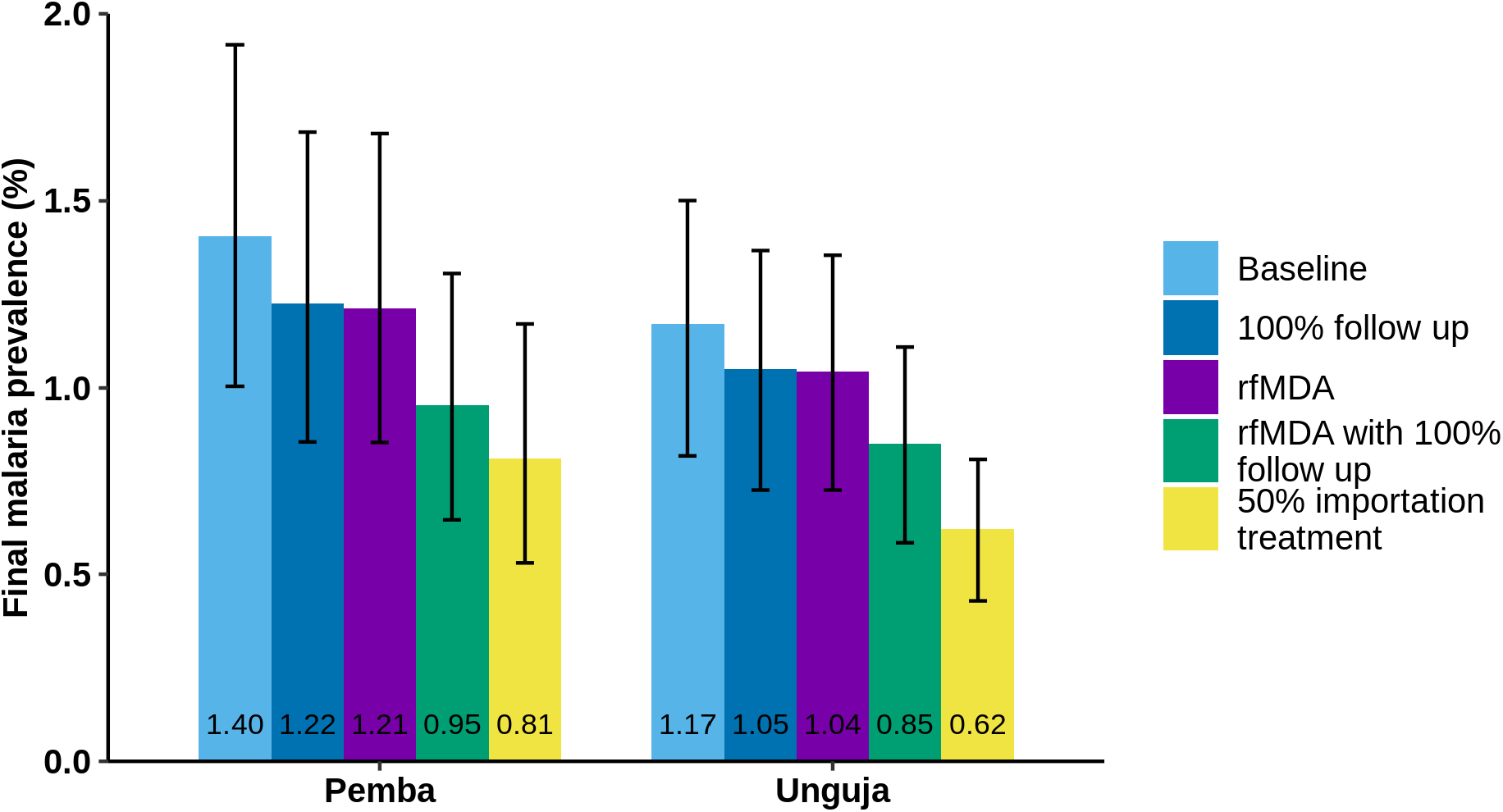
Bar chart showing the final equilibrium value after 40 years of the SIS model with the current RCD system (baseline), an RCD system with 100% follow up, replacing the RCD system with rfMDA, both replacing the RCD system with rfMDA and increasing follow up to 100%, and treating 50% of imported cases while maintaining the baseline RCD programme. The error bars here show the 95% confidence interval for both the stochastic variation and parameter uncertainty.

## 4 Discussion

Our results suggest that case importation and the low test sensitivity of RDTs in asymptomatic patients are the main factors that should be targeted to substantially reduce Zanzibar’s malaria burden, while continuing to maintain the vector control measures that are currently in place. Removing the RCD programme would likely lead to an increase in malaria prevalence, but increasing follow up to cover all malaria cases arriving at a health facility would still be insufficient for reaching elimination. Treating imported cases, implementing rfMDA at the household level and increasing the rate at which infected people seek treatment would help reduce the endemic prevalence on both islands substantially. 100% imported case treatment is expected to reduce the prevalence below 1 case per 100,000 on Unguja and 1.4 cases per 10,000 population on Pemba, as Zanzibar acts as a sink for infections from mainland Tanzania, where prevalence is higher. This result assumes that all current measures are maintained. Relaxing interventions already in place may lead to the local reproduction number being higher than 1, and thus elimination would not be achieved even with treating 100% of imported cases.

As those residing in the same household as index cases are significantly more likely to test positive for malaria than those residing in neighboring households [3], the extra effort of testing neighboring residents makes little difference to overall transmission as compared to increasing follow up at the households of index cases. Expanding RCD to include neighbours requires extra resources and the reduction in malaria prevalence is minimal in comparison to the extra RDTs and human resources required. Nonetheless, surveillance is a key component of establishing when malaria elimination has occurred, so some form of passive or active surveillance is required to monitor cases. This can be RCD or just case reporting, but RCD allows for the surveillance of asymptomatic cases as well.

Moving from RCD to rfMDA allows for the treatment of approximately three times more cases for any given prevalence, particularly low density infections that are less likely to be detected by RDT, but may still contribute to onward transmission. It is possible that early infections in neighbours are missed by RCD as the parasite density may be too low to be detected by RDTs. A previous field study compared RCD to rfMDA in the low malaria-endemic setting of Namibia and found a significant reduction in incidence in the rfMDA arm [16]. rfMDA in this context could also have a prophylactic effect, preventing onward transmission from the index case. However, rfMDA involves substantially greater use of ACTs than RCD. This may have a negative effect on parasite resistance [34, 35]. Increases in drug resistance may lead to increased treatment failure rates, leading to a resurgence in malaria prevalence, though this was not found to be a frequent cause of malaria resurgence in previous work [36].

These results are broadly in line with findings from other studies on RCD effectiveness in different settings. A recent study of mass drug administration campaigns in the Greater Mekong Subregion suggested that an RCD programme in the region would have missed 99.6% of *Plasmodium* infections [37]. When modelling RCD in southern Zambia, the number of people presenting to a health facility with malaria and being followed up was found to be a limiting factor for an RCD programme’s success [13]. Similarly, an independent study of Zambia’s reactive case detection system found that in low-transmission settings, improving case management (the rate at which patients seek treatment from health facilities) would have a greater impact on onward transmission than further improving the RCD system [14]. Additionally, this study highlighted that in both low and high transmission settings, importation management was crucial for successful disease elimination. Similar results were found by Le Menach *et al* when examining malaria importation rates onto Pemba and Unguja in 2012 [18]. Our findings also show that importation management is key to interrupting transmission on Unguja and substantially reducing disease prevalence on Pemba. As the average time spent on mainland Tanzania is higher amongst Unguja residents in the sample, as compared to Pemba residents, the effect of importation was estimated to be larger on Unguja than on Pemba.

We calibrate the transmission rate based on the malaria prevalence in the three patches and the movement between the patches. We make the simplifying assumption that factors such as immunity profiles, healthcare access, and the proportions of patients who are asymptomatic are identical amongst travellers and non-travellers due to a lack of empirical data from Tanzania on these factors. This may not necessarily be true, as in some areas, migration is associated with less use of healthcare facilities and higher malaria risk profiles [38]. This would suggest that malaria in travellers might play a larger role in transmission than described in this study. On the other hand, repeated exposure to malaria among travellers might lead them to have greater levels of immunity than non-travellers, and so they may have lower parasite densities and, subsequently, lower infectiousness when infected. In that case, the impact of case importation may be smaller than described. We also assume that longer-term migrants do not play a significant role in case importation in Zanzibar. This is supported by a study looking at census data and malaria transmission rates in East Africa, which suggests that most migrants from high transmission areas settle near the borders of mainland Tanzania, but not many come to Zanzibar [25].

Reconstructing travel history data from survey responses is prone to underestimates of travel frequency, as certain trips may not be recalled. Thus, our estimate of the amount of time Zanzibari residents spend away from home are likely to be underestimates. Therefore, malaria importation is likely to play a larger role in malaria persistence than estimated here. We have also not considered the seasonal variation in travel. The busiest travel period typically falls between October and December, which coincides with the shorter period of seasonal rainfall [2, 18]. This variation throughout the year will also impact the rate of case importation into the region.

As RDTs typically detect cases with a higher parasite density, and cases with a higher parasite density are more likely to be symptomatic, RCD may, over time, lead to the infectious reservoir being skewed towards asymptomatic infections. In this model, we model all infections as having equal infectiousness, whereas these asymptomatic infections may have a lower infectiousness, and so the impact of RCD may be greater than that displayed here.

Additionally, we have assumed that malaria transmission is constant throughout the year on the islands. The data used in this study is averaged across both high and low seasons of transmission [3]. Seasonal transmission likely increases the importance of imported cases, as elimination may be achieved in the dry season, but cases are re-introduced in the wet season when the transmission rate is higher. Also, reactive vector control is another reactive intervention that may be considered in the wet season, which would involve spraying insecticide inside index and neighbouring households to prevent further transmission from known cases. A field study of reactive vector control found adding it to RCD or rfMDA had an additional benefit in reducing malaria incidence in Namibia [16].

This analysis does not preclude the existence of smaller foci of transmission that could exist on these islands. Transmission is likely to be heterogeneous, with local sources and sinks of cases. As there was insufficient data on local movement patterns within each island, each island has been treated as homogeneous. Extending this analysis with other sources of data on travel, such as call record detail data, would allow for a finer-scale analysis of parasite sources and sinks.

Additionally, as the model presented here is an SIS model, it does not include the relationship between infection and disease, which would play a role in the effectiveness of an RCD programme that relies on patients seeking treatment. This should be considered in future work conducted in this area.

Here, we have defined malaria elimination as having zero malaria infections present on an island. In contrast, the World Health Organization defines a country to have eliminated malaria when they have zero indigenous cases for three consecutive years, allowing for some imported and introduced cases [1]. Thus, our definition of elimination is a stricter definition in comparison to the World Health Organization.

## 5 Conclusion

Our analysis suggests that the current interventions in place on Unguja have sufficiently reduced the transmission rate such that malaria elimination could be achieved in the absence of imported cases. On Pemba, the situation is less clear, though the mean controlled reproduction number is below 1. Current interventions should be maintained, and improvements to the surveillance-response system are expected to have an incremental effect on the malaria prevalence. Interventions with the most impact were found to be those that removed the majority of cases imported to the islands.

## Data Availability

All data used in this study are available online at https://github.com/SwissTPH/ZanzibarRCD

## Acknowledgements

We would like to thank Thomas Smith, Monica Golumbeanu and Pascal Grobecker for their helpful discussions and feedback on this manuscript.

## Funding

NC and AMD were supported by the Bill and Melinda Gates Foundation (OPP1032350 and INV025569). Funding for the RADZEC study was provided by the Swiss Tropical and Public Health Institute and the US President’s Malaria Initiative via the US Agency for International Development/Tanzania under the terms of an inter-agency agreement with Centers for Disease Control and Prevention (CDC) and the US Agency for International Development/Tanzania through a cooperative agreement with the MEASURE Evaluation consortium, under the associate cooperative agreement No. AID-621-LA-14-00001 titled ‘Measure Phase III— Strengthening the monitoring, evaluation and research capacity of the community health and social service programmes in the United Republic of Tanzania’. The opinions expressed herein are those of the authors and do not necessarily reflect the views of the President’s Malaria Initiative via the US Agency for International Development, or other employing organizations or sources of funding.

## Authors’ contributions

AMD contributed to the conceptualization, methodology, formal analysis and writing of this manuscript; MAH and JOY contributed to the conceptualization and critically reviewed the manuscript; LS contributed to the investigation, data curation and critically reviewed the manuscript; BSF, AHA and AA contributed to the investigation and critically reviewed the manuscript; NC contributed to the conceptualization, methodology and supervision of the study, and critically reviewed the manuscript. All authors gave final approval for publication and agree to be held accountable for the work performed therein.

## Notes

### Competing Interest Statement

The authors have declared no competing interest.

### Funding Statement

NC and AMD were supported by the Bill and Melinda Gates Foundation (OPP1032350). Funding for the RADZEC study was provided by the Swiss Tropical and Public Health Institute and the US President's Malaria Initiative via the US Agency for International Development/Tanzania under the terms of an inter-agency agreement with Centers for Disease Control and Prevention (CDC) and the US Agency for International Development/Tanzania through a cooperative agreement with the MEASURE Evaluation consortium, under the associate cooperative agreement No. AID-621-LA-14-00001 titled *Measure Phase III-- Strengthening the monitoring, evaluation and research capacity of the community health and social service programmes in the United Republic of Tanzania*. The opinions expressed herein are those of the authors and do not necessarily reflect the views of the President's Malaria Initiative via the US Agency for International Development, or other employing organizations or sources of funding.

### Author Declarations

Ethical approval for the collection of the data used in this study was obtained from the Zanzibar Medical Research Ethics Committee, the Institutional Review Boards of Tulane University and of the Ifakara Health Institute as well as the Ethics Commission of North-western and Central Switzerland.

### Summary of Updates

Paper title updated. Figures have been updated, results remain unchanged. Additional analysis of ACT and RDT use for the different RCD-related interventions is now included. The discussion has been expanded to consider some other limitations of this study and the potential impact on results.

