## Supplementary Information for "The impact of reactive case detection on malaria transmission in Zanzibar in the presence of human mobility"

### S1 Methods

#### S1.1 The controlled reproduction number for the whole system

The controlled reproduction number for the whole system, $R_s$, gives the expected number of secondary infections arising across all three patches from a primary infection when interventions are in place. $R_s$ was calculated by taking the spectral radius of the next generation matrix [1, 2]. Note, $R_s$ is different to the local controlled reproductive number on each island given in the main text, $R_c$, as that is calculated by taking the ratio of the transmission and recovery rates on each island.

The equation for the rate of change of infected individuals (equation (4) in the main text) can be linearised by decomposing the Jacobian matrix into two matrices describing the transmission events leading to new infections, $\mathbf{F}$, and the recovery events leading to removal from the infected class, $\mathbf{V}$:

$$\frac{\mathrm{d}\vec{I}}{\mathrm{d}t} = (\mathbf{F} - \mathbf{V})\vec{I}, \tag{1}$$

where

$$F_{ij} = \sum_{k=1}^{3} \left( \frac{\beta_k N_j \theta_{kj} \theta_{ki}}{\sum_{l=1}^{3} N_l \theta_{kl}} \right), \tag{2}$$

[*]Corresponding author
[†]Current affiliations: Amsterdam Institute for Global Health and Development, Amsterdam, Netherlands and Amsterdam University Medical Centers, Amsterdam, Netherlands
[‡]Current affiliation: RTI International, Dar es Salaam, United Republic of Tanzania

and

$$V_{ij} = \begin{cases} \mu & i = j, \\ 0 & i \neq j. \end{cases} \tag{3}$$

The next generation matrix, $\mathbf{K}$, is given by

$$\mathbf{K} = \mathbf{F}\mathbf{V}^{-1}. \tag{4}$$

$R_s$ is equal to the spectral radius of $\mathbf{K}$.

In order to investigate the potential impact of changing transmission levels on mainland Tanzania, the value of $\beta_{\text{mainland}}$ was varied between 0.004 and 0.0054 and $R_s$ was re-calculated.

The results are shown in Fig. S1. While the value for the transmission rate on mainland Tanzania is below the transmission rate on Pemba, the spectral radius of $\mathbf{K}$ is dominated by the ratio of the transmission and recovery rates on Pemba. Once the transmission rate on the mainland approaches that of Pemba, the mainland transmission rate becomes the dominant factor in determining the spectral radius of $\mathbf{K}$. After this point, we see a linearly increasing relationship between the mainland transmission rate and the controlled reproductive number of the system.

##### S1.2 Treatment of imported cases

Treatment of imported cases leads to a shift in the equilibrium prevalence observed on each island. Fig. S2 shows the timeseries plots for reaching equilibrium for the following proportions of treatment of outbound travellers from the mainland to Zanzibar, $O$, and travellers from Zanzibar on their return to Zanzibar, $R$, assuming the baseline level of RCD is maintained:

- $O = R = 0.25$,
- $O = R = 0.5$,
- $O = R = 0.75$,
- $O = R = 1$.

#### S2 Figures

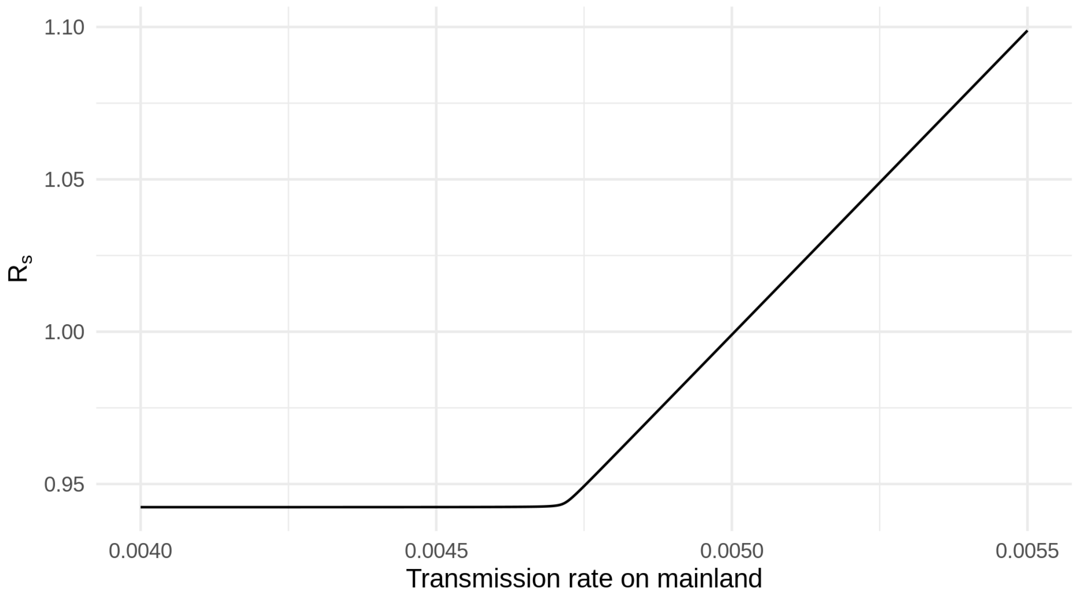


**Figure S1:** Plot showing the relationship between the transmission rate on mainland Tanzania, and the controlled reproductive number of the whole system.

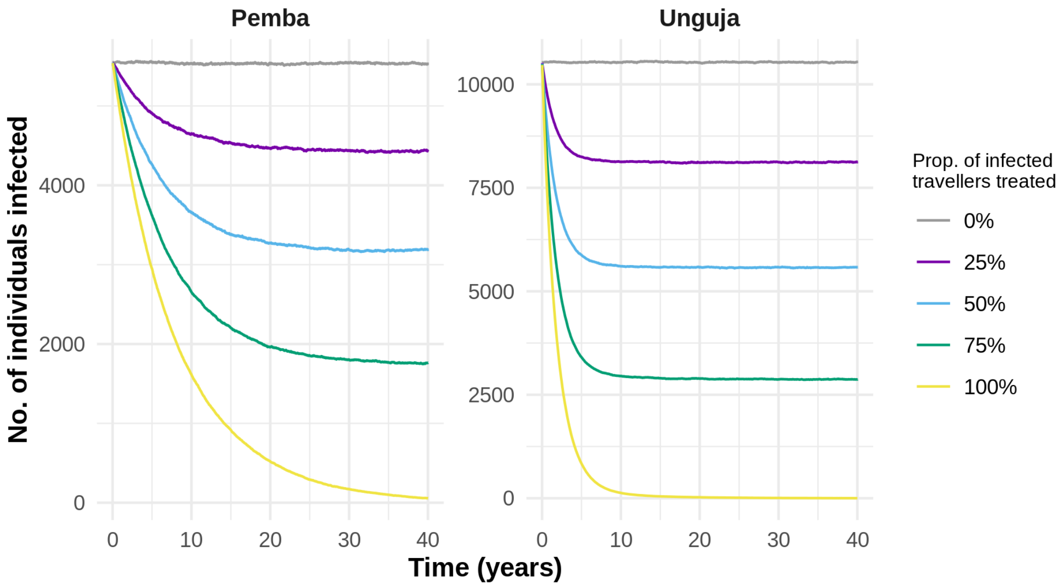


**Figure S2:** 7-day moving average of the median of 500 stochastic simulations for an SIS model of RCD for Pemba and Unguja comparing different levels of treating infected travellers, assuming baseline RCD is maintained. Here, the value given as the proportion of travellers treated account for both travellers from the mainland and travellers from Zanzibar.
